# Trends in Why Americans Delayed Dental Care From Pre-Pandemic to the COVID-19 Era: Implications for Aging and Oral Public Health

**DOI:** 10.64898/2026.08.22.26361060

**Authors:** Preeti Pushpalata Zanwar, Jay Sureshbhai Patel, Chan Shen

## Abstract

**Objectives:** To describe age-group differences in inability to afford dental treatment and cost related dental delay, among the US community-dwelling population.

**Study design:** Descriptive analysis of nationally representative survey data.

**Methods:** Using nationally representative Medical Expenditure Panel Survey data (2018-2021), we examined trends in inability to afford dental treatment and cost-related dental treatment delays across four age groups (2-17, 18-39, 40-64, ≥65 years). Weighted analyses accounted for the complex survey design; statistical significance was set at *p*<0.001.

**Results:** Cost-related delays declined modestly from 2018 to 2021 but remained most prevalent among adults aged 40-64 (4.8% for ages 40-64, 3.4% for ages 18-64, 2.2% for ages≥65 in 2021; p<0.001).

**Conclusion:** Middle-aged adults seem to experience delays due to cost, underscoring the need for dental coverage to expand dental coverage for this group and to reduce their out-of-pocket costs.

## INTRODUCTION

Oral health and systemic health share a well-established bi-directional relationship. Oral diseases such as periodontitis are progressive in nature and associated with chronic systemic illnesses including diabetes, cardiovascular disease, and adverse pregnancy outcomes, while systemic diseases can, in turn, exacerbate oral disease progression.^1^ Timely dental care is essential for preserving oral function and preventing disease progression, especially for middle-aged and older adults, who may have other co-occurring chronic conditions; delays can worsen oral conditions, increase complexity and cost, and contribute to poor oral and systemic health and QOL.^2-3^

Dental care coverage and dental benefits vary widely by age group.^4^ For older adults, in the United States (U.S.), the traditional Medicare government-sponsored health insurance program does not offer dental benefits, while working-age middle-aged adults often lack comprehensive dental benefits.^4-5^ Middle-aged adults are disproportionately affected because of insurance gaps and high OOP costs, highlighting affordability as a major barrier to dental care even when services are physically available and a key determinant of oral health outcomes.^2,2-2^ Dental care therefore presents higher financial barriers than most other health services, largely due to high out-of-pocket (OOP) costs.^2,2-2^ Moreover, the COVID-19 pandemic represented an unprecedented disruption to health care delivery and to the delivery of non-urgent dental services.

During the early phases of the pandemic, many dental practices were temporarily closed or limited to emergency-only care due to shutdown orders, infection control concerns, and workforce constraints.^2-3,8^ Even as practices reopened, fear of infection and restricted services further limited utilization.^8^ These pandemic-related disruptions compounded existing affordability barriers and resulted in widespread forgone dental care.^2-3^ Prior research has identified insurance status and financial barriers as key determinants of dental care utilization.^4-5,8^

However, much of the existing literature has assessed the effects of large-scale national disruptions such as the COVID-19 pandemic.^2-4^ Limited studies have simultaneously assessed age heterogeneity across the lifespan among community-living Americans in cost- and affordability barriers pre-COVID-19 and alongside COVID-19-related disruption.

Evaluating financial barriers patterns amidst pandemic-related disruptions over time allows for understanding whether barriers to care were temporary or persistent before the pandemic, and whether pandemic-related disruptions intensified age-based disparities in reasons for dental delays. Such analyses are particularly important across different age groups, which vary substantially in insurance coverage, employment stability, and dental care access.^4^ Our purpose was to estimate the heterogeneity for the reasons for delays in dental treatment before during COVID-19 using nationally representative data on individuals residing in U.S. community living households. Specifically, we sought to compare pre-pandemic and pandemic-era patterns in affordability and cost-related differences in dental care treatment across four mutually exclusive age groups.

## METHODS

### Data Source

We leveraged the nationally representative Medical Expenditure Panel Survey (MEPS) Household Component individual-level data available from https://meps.ahrq.gov/mepsweb/ on non-institutionalized community-living persons [9]. Children and adults residing in the United States; data collected on Panel 23 using 4-year file spanning years 2018 through 2021.^9^ Conceptually, pre-pandemic was designated as years 2018-2019, while COVID-19 as years 2020-2021. Given that MEPS data are de-identified and publicly available, our study was exempt from Thomas Jefferson University’s Institutional Review Board.

### Primary Measures

Using the access to care supplement questions, we estimated for each year the proportion of individuals across four age groups (ages 2-17, 18-39, 40-64, ≥65 years using a pre-constructed age variable reported that captured a person’s age in December for each particular year) whether a person was delayed in receiving dental care: 1) because they could not afford it, and 2) whether they worried about cost.

### Statistical Analysis

We accounted for the MEPS complex survey design, including stratification, clustering, and sampling by including primary sampling unit, variance estimation, and person weights to generate weighted proportions of nationally representative estimates of our primary measures cross-sectionally across the four years. We assessed differences in proportions across the four age groups separately for each year using the F-test. Statistical significance was defined as a two-sided p-value <0.001. All analyses were conducted using Stata 16.

## RESULTS

Annual unweighted sample sizes of our analytic sample varied for each year, ranging from 6503 to 6536 unweighted, representing more than 310 million US residents.

Figure 1 (Panels A-C) and Supplementary Tables 1-3 (Panels A-C) show that the proportion of individuals unable to afford dental care declined from 2018 to 2021 (overall 15.6% in 2018 to 10.8% in 2021). Age was strongly associated with affordability in all years (p<0.001), with adults aged 40-64.9 years consistently reporting the highest prevalence of unaffordability (5.4% in 2018, 5% in 2019; 3.7% in 2020 and 3% in 2021). Adults aged 18-39.9 years also reported more affordability barriers than children and older adults (in 2020, 5.1% among ages 18-39.9 years vs.1.9% among ages ≥65 years, p<0.001; in 2021, 3.4% among ages 18-39.9 years vs. 2.2% among ages ≥65 years, p<0.001), these patterns are consistent with known differences in dental coverage and expenses [6-9]. Among middle-aged adults 40-64.9 years, cost-related delays were common before COVID-19 and declined modestly during the pandemic years (in 2019, 6.5% vs. 4.8% among ages 40-64.9 years), see Figure 1, Panel B, Supplementary Table 2).

**Figure 1.**
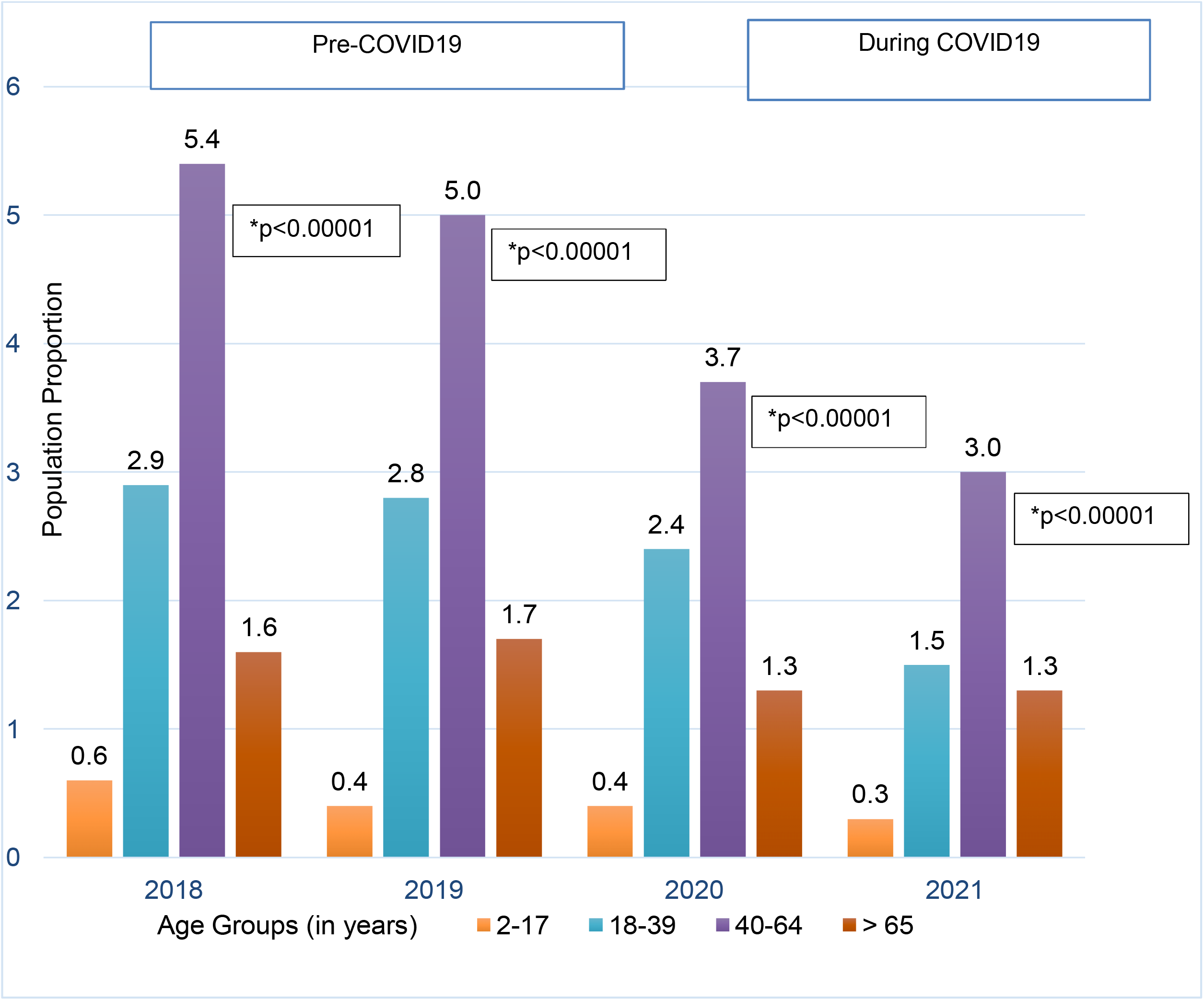
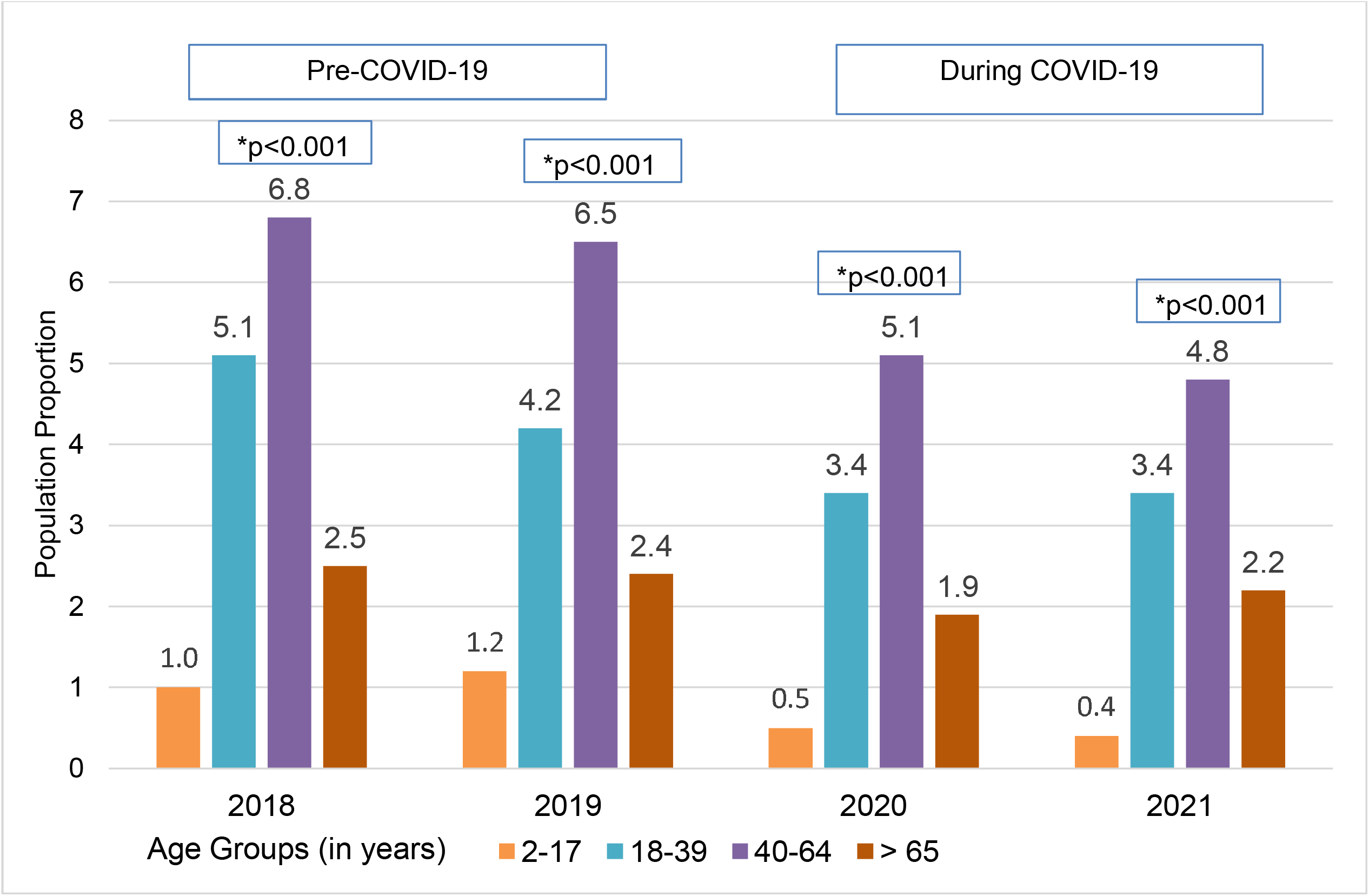
**(Panel A):** Weighted U.S. Proportion by Age Group that Reported Delays in Dental Care Because Could Not Afford Dental Treatment, Medical Expenditure Panel Survey, Years 2018-2021.

## DISCUSSION

Affordability barriers remained common and strongly patterned by age, with middle-aged adults experiencing the greatest burden. Adults aged 18-39.9 years reported more affordability barriers than children and older adults, which is consistent with known differences in patterns of dental coverage and OOP exposure.^2-7^ In our study during 2020-2021, reports of unaffordability may have been due to several reasons, such as insurance changes or shifts in perceived need.^4-7^

Across all years, age was strongly associated with cost-related delays, with middle-aged adults reporting the highest prevalence, followed by younger adults. These findings reinforce prior evidence that cost-related access barriers are structurally patterned by age and insurance coverage.^4-5^ The decline observed during 2020–2021 likely reflects pandemic-related disruptions in routine care rather than resolution of financial barriers.^2-4^ The patterns observed in our study seem to be consistent with the reopening of dental practices, when strengthening of infection-control protocols, and vaccine rollout occurred.^8^ In contrast, age showed a strong age gradient for cost-related barriers.

### Implications for Aging and Oral Public Health

Delays in dental treatment are central to disease prevention, equity, and dental policy.^2-4^ First, dental treatment delays 1) transform preventable oral disease into more severe, costly, and unequal health burdens across the population.^2-4^ Second, they increase rates of caries, periodontal disease, systemic health issues, tooth loss, pain, social isolation, and infection, which in turn worsen nutrition, speech, employability, and overall quality-of-life.^1^ Third, they drive up treatment costs by shifting care from prevention to emergency and complex procedures, straining both public programs and household finances. Finally, they exacerbate health disparities as low-income uninsured, and working-age adults are more likely to delay care for cost or access reasons, leading to concentrated unmet need.^2-3,5^ During COVID-19, widespread postponement of routine dental visits amplified these risks.^2^

### Limitations

We report cross-sectional patterns in dental delays during 2018 and 2021, and causality between the pandemic and delays cannot be inferred. We cannot differentiate any pandemic-related mechanisms, such as social distancing or closures, for which MEPS does not collect data, but we acknowledge that these can bias findings. While response rates for MEPS declined during COVID-19, MEPS data collected during this time are robust and validated for use.^10^ MEPS data collection response rate considerably dropped during the pandemic due to a halt in face-to-face interviews but did not affect the quality of data collected.^10^ The current study used a unique 4-year MEPS data file. The overall lower response rates and increased nonresponse bias across subsequent data collection rounds may have reduced the statistical precision of estimates.^10^ We acknowledge that older adults on Medicare may be delaying dental treatment for other reasons than cost, such as mobility problems, transportation barriers, and fear and anxiety.^5^

Public policy solutions for pandemic-resilient dental systems can include policies that place importance on affordable dental care, providing continuous access, and integration of medical-dental delivery models with an emphasis on prevention and early intervention. Structural reforms aimed at reducing OOP costs, particularly for middle-aged adults. This may be one of the solutions to improving timely dental treatments in oral public health.

## Data Availability

MEPS data are publicly avaialble from https://meps.ahrq.gov/mepsweb/

## Conflicts of Interest: Conflicts of Interest

PPZ serves as the Group Program Chair for the Geriatric Oral Research Group (GORG) at the International Association of Dental, Oral, and Craniofacial Research (IADR)

### Use of any Artificial Intelligence Generated Content (AIGC) tools

No AI tool was used to design the study, for data construction, data or statistical analysis, or for any figures or for generating references. Academic Perplexity was used to assess any discrepancies in the introduction and discussion, which were carefully edited, reviewed, and revised by all co-authors. The corresponding author carefully read and revised all content and takes full responsibility for the article’s originality and integrity.

## Appendix A

**Supplementary Table 1.**
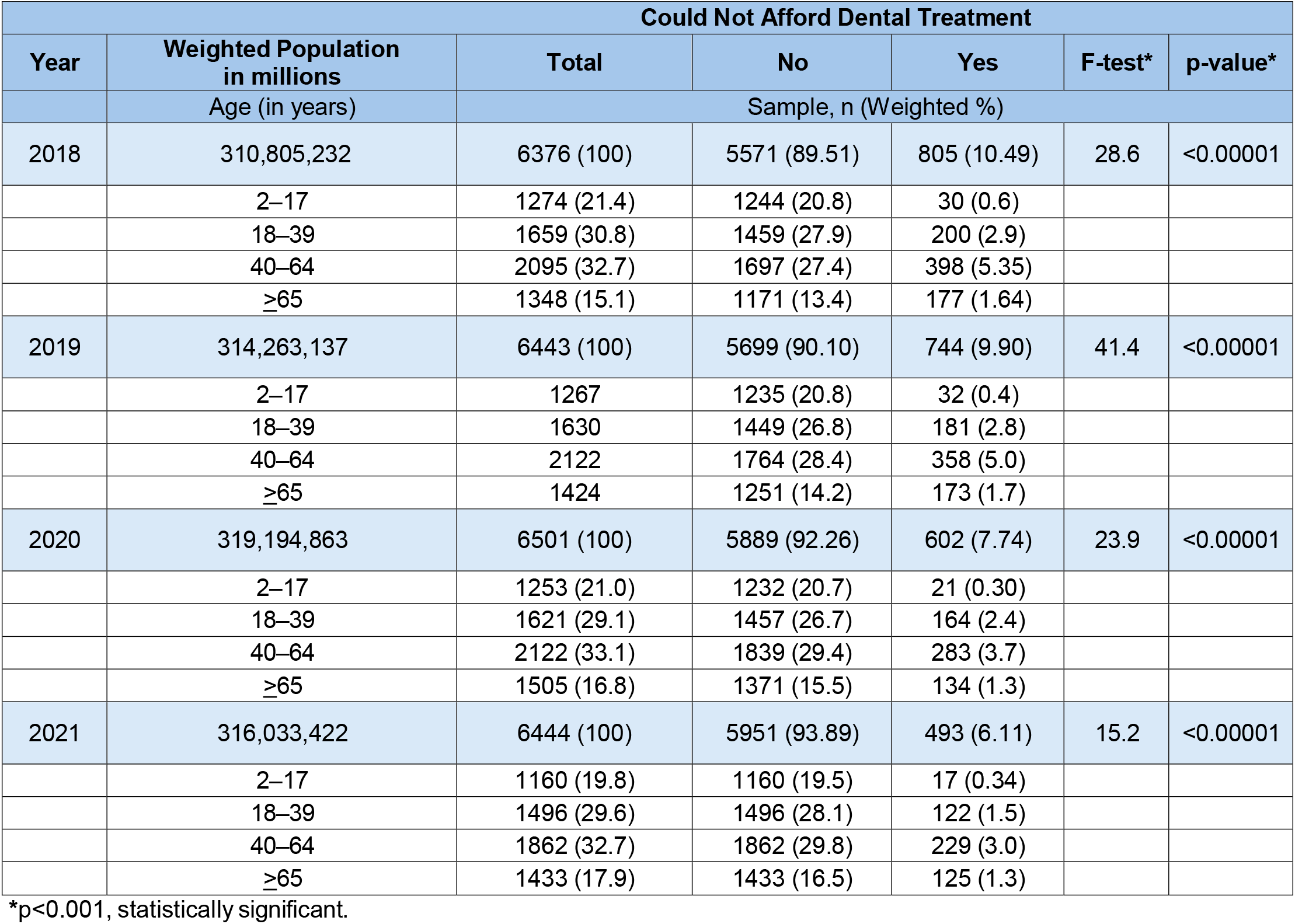
**(Figure 1: Panel A):** Population Characteristics that Could Not Afford Dental Treatment, Medical Expenditure Panel Survey, Years 2018-2021.

**Supplementary Table 2.** **(Figure 1: Panel B):** Population Characteristics Who Reported Delays in Dental Treatment Because of Cost, Medical Expenditure Panel Survey, Years 2018-2021.

| Year | Overall Population<br>(Weighted, in millions) | Delays in Dental Treatment Because of Cost |  |  |  |  |
| --- | --- | --- | --- | --- | --- | --- |
|  |  | Total | No | Yes | F-test* | p-value* |
|  | Age (in years) | Sample, n (Weighted %) |  |  |  |  |
| 2018 | 310,963,500 | 6377(100) | 5280 (84.5) | 1097 (15.5) | 25.3 | <0.001 |
|  | 2-17 | 1276 (21.4) | 1221 (20.4) | 55 (1.0) |  |  |
|  | 18-39 | 1660 (30.8) | 1351 (25.6) | 309 (5.1) |  |  |
|  | 40-64 | 2096 (32.8) | 1610 (25.9) | 486 (6.8) |  |  |
|  | ≥65 | 1276 (15.0) | 1098 (12.6) | 247 (2.5) |  |  |
| 2019 | 314,381,356 | 6443 (100) | 5419 (85.7) | 1024 (14.3) | 25.6 | <0.001 |
|  | 2-17 | 1267 (21.2) | 1195 (20.0) | 72(1.2) |  |  |
|  | 18-39 | 1630 (29.6) | 1377 (25.4) | 253 (4.2) |  |  |
|  | 40-64 | 2124 (33.4) | 1665 (26.9) | 459 (6.5) |  |  |
|  | ≥65 | 1422 (15.9) | 1182 (13.5) | 240 (2.4) |  |  |
| 2020 | 319,371,163 | 6507 (100) | 5706 (89.2) | 801 (10.6) | 24 | <0.001 |
|  | 2-17 | 1253 (21.0) | 1223 (20.5) | 30 (0.5) |  |  |
|  | 18-39 | 1624 (29.1) | 1407 (25.7) | 217 (3.4) |  |  |
|  | 40-64 | 2124 (33.1) | 1755 (28.1) | 369 (5.1) |  |  |
|  | ≥65 | 1506 (16.8) | 1321 (14.9) | 185 (1.9) |  |  |
| 2021 | 316,129,606 | 6451 (100) | 5655 (89.2) | 796(10.8) | 23.6 | <0.001 |
|  | 2-17 | 1178 (19.8) | 1151 (19.4) | 27(0.4) |  |  |
|  | 18-39 | 1619 (29.6) | 1410 (26.2) | 209 (3.4) |  |  |
|  | 40-64 | 2094 (32.7) | 1735 (27.9) | 359 (4.8) |  |  |
|  | ≥65 | 1560 (17.9) | 1359 (15.7) | 201 (2.1) |  |  |

\*p<0.001, statistically significant.

